# Genomic Diversity of SARS-CoV-2 During Early Introduction into the United States National Capital Region

**DOI:** 10.1101/2020.08.13.20174136

**Authors:** Peter M. Thielen, Shirlee Wohl, Thomas Mehoke, Srividya Ramakrishnan, Melanie Kirsche, Oluwaseun Falade-Nwulia, Nídia S. Trovão, Amanda Ernlund, Craig Howser, Norah Sadowski, Paul Morris, Mark Hopkins, Matthew Schwartz, Yunfan Fan, Victoria Gniazdowski, Justin Lessler, Lauren Sauer, Michael C. Schatz, Jared D. Evans, Stuart C. Ray, Winston Timp, Heba H. Mostafa

**Affiliations:** Johns Hopkins University Applied Physics Laboratory, Laurel, Maryland 20723; Johns Hopkins Bloomberg School of Public Health, Baltimore, Maryland 21205; Johns Hopkins University Department of Computer Science, Baltimore, Maryland 21218; Johns Hopkins University School of Medicine, Department of Medicine, Division of Infectious Disease, Baltimore, Maryland 21205; National Institutes of Health, Fogarty International Center, Bethesda, Maryland 20892; Johns Hopkins University School of Medicine, Department of Pathology, Division of Medical Microbiology, Baltimore, Maryland 21205; Johns Hopkins University Department of Emergency Medicine, Baltimore, Maryland 21205; Johns Hopkins University Departments of Biomedical Engineering and Molecular Biology and Genetics, Baltimore, Maryland 21205

## Abstract

**Background:** The early COVID-19 pandemic has been characterized by rapid global spread. In the United States National Capital Region, over 2,000 cases were reported within three weeks of its first detection in March 2020. We aimed to use genomic sequencing to understand the initial spread of SARS-CoV-2, the virus that causes COVID-19, in the region. By correlating genetic information to disease phenotype, we also aimed to gain insight into any correlation between viral genotype and case severity or transmissibility.

**Methods:** We performed whole genome sequencing of clinical SARS-CoV-2 samples collected in March 2020 by the Johns Hopkins Health System. We analyzed these regional SARS-CoV-2 genomes alongside detailed clinical metadata and the global phylogeny to understand early establishment of the virus within the region.

**Results:** We analyzed 620 samples from the Johns Hopkins Health System collected between March 11–31, 2020, comprising 37.3% of the total cases in Maryland during this period. We selected 143 of these samples for sequencing, generating 114 complete viral genomes. These genomes belong to all five major Nextstrain-defined clades, suggesting multiple introductions into the region and underscoring the diversity of the regional epidemic. We also found that clinically severe cases had genomes belonging to all of these clades.

**Conclusions:** We established a pipeline for SARS-CoV-2 sequencing within the Johns Hopkins Health system, which enabled us to capture the significant viral diversity present in the region as early as March 2020. Efforts to control local spread of the virus were likely confounded by the number of introductions into the region early in the epidemic and interconnectedness of the region as a whole.

## Introduction

Within two months of its emergence in Wuhan, China in early 2020, SARS-CoV-2, the virus that causes COVID-19, had established itself worldwide^1^: as of July 30, 2020, over 10 million confirmed COVID-19 cases and 650,000 deaths have been reported^2^. The enormous health and economic impacts of this virus have led to considerable interest into understanding its origin, spread, and evolution. Generation and analysis of pathogen genomic data has been a key component of this research^3-8^ and promises to provide critical insights into not only the emergence and spread of SARS-CoV-2, but the dynamics of emerging infections in general.

In the United States, diagnostic capacity for SARS-CoV-2 was limited until early March 2020 due to regulatory challenges associated with limited Emergency Use Authorization (EUA) for laboratory-developed testing. Retrospective analyses of patient samples using genomic and serological methods now suggests that community transmission was occuring in major US cities in late January and early February of that year^9-11^. Ongoing work has continued to deepen our understanding of SARS-CoV-2, including use of pathogen sequence data to help reconstruct the transmission of the virus into and around the United States^12^.

Coronaviruses, including SARS-CoV-2, possess an RNA polymerase with proofreading activity that limits genetic variability ^13^. This genome replication feature, combined with rapid spread and limited or no host immunity during the early phase of the pandemic, has likely limited evolutionary pressure and contributed to the limited genetic diversity observed in SARS-CoV-2 sequences. Efforts to describe this diversity during the early phase of the pandemic have resulted in multiple clade designation systems including those used by the Global Information Sharing of All Influenza Data (GISAID), the NextStrain platform, and COVID-19 Genomics UK consortium (COG-UK)^14-16^. These approaches are intended to be dynamic, and are updated as new diversity is observed in the global sequence data. For example, NextStrain clade designations are broadly separated by viruses that originated in China in 2019 (Clade 19) and those that were later introduced into Europe early 2020 (Clade 20)^15^. Importantly, SARS-CoV-2 clade designations are only intended to identify subgroups of virus sequences that share common genetic features, and further *in vitro* or clinical characterization is required to identify functional differences between clades.

Using sequence data to investigate relationships between virus genetics and patient clinical outcome often relies on specimen repositories or agreements with sample collection facilities that provide limited access to patient demographic and clinical information. Limited access is due to both logistical challenges in obtaining this data and ethical concerns around patient privacy^17^. Therefore, while research efforts have produced copious amounts of valuable genetic data and insights into viral circulation ^12,18^, studies linking pathogen genetics to demographics, disease severity, and other clinical outcomes are less frequent. By leveraging existing internal networks and established research protocols, research groups within large health systems can fill this gap by rapidly creating and analyzing datasets that link pathogen genomics with clinical and demographic outcomes.

To gain insight into regional viral spread and potential associations between pathogen genetics and clinical outcomes, we performed rapid whole genome sequencing of the SARS-CoV-2 virus from clinical samples in the Johns Hopkins Health System (JHHS). For this work we used the Oxford Nanopore sequencing platform, which has been increasingly used during early outbreak investigations to understand emerging pathogens^19,20^. In the process of establishing SARS-CoV-2 sequencing capacity within the JHHS, we developed a number of data analysis improvements that increase confidence in identifying single nucleotide polymorphisms from Oxford Nanopore sequencing data. Here, we explore the relationship between local sequences and those from the broader national and global epidemic, and look for possible associations between clade structure and clinical outcomes.

## Results

### Characteristics of SARS-CoV-2 Identification in the Region

The Maryland, District of Columbia (DC), and Virginia area is designated as the United States National Capital Region — an area of high domestic and international transit as well as geopolitical importance. The Johns Hopkins Health System (JHHS) is spread throughout the region, including 39 hospitals and clinics throughout Maryland and DC, with a patient population that also includes residents of Northern Virginia. The first detection of SARS-CoV-2 in the region was reported on March 5, 2020 by the Maryland Department of Health, and a State of Emergency was immediately declared in Maryland ^21^. Two days later cases were reported by the Public Health Departments in Virginia and Washington DC. On March 23^rd^, the Maryland state government instituted regional closure of non-essential businesses, and a stay at home order followed on March 30^th^ (**Fig 1A**).

**Figure 1:**
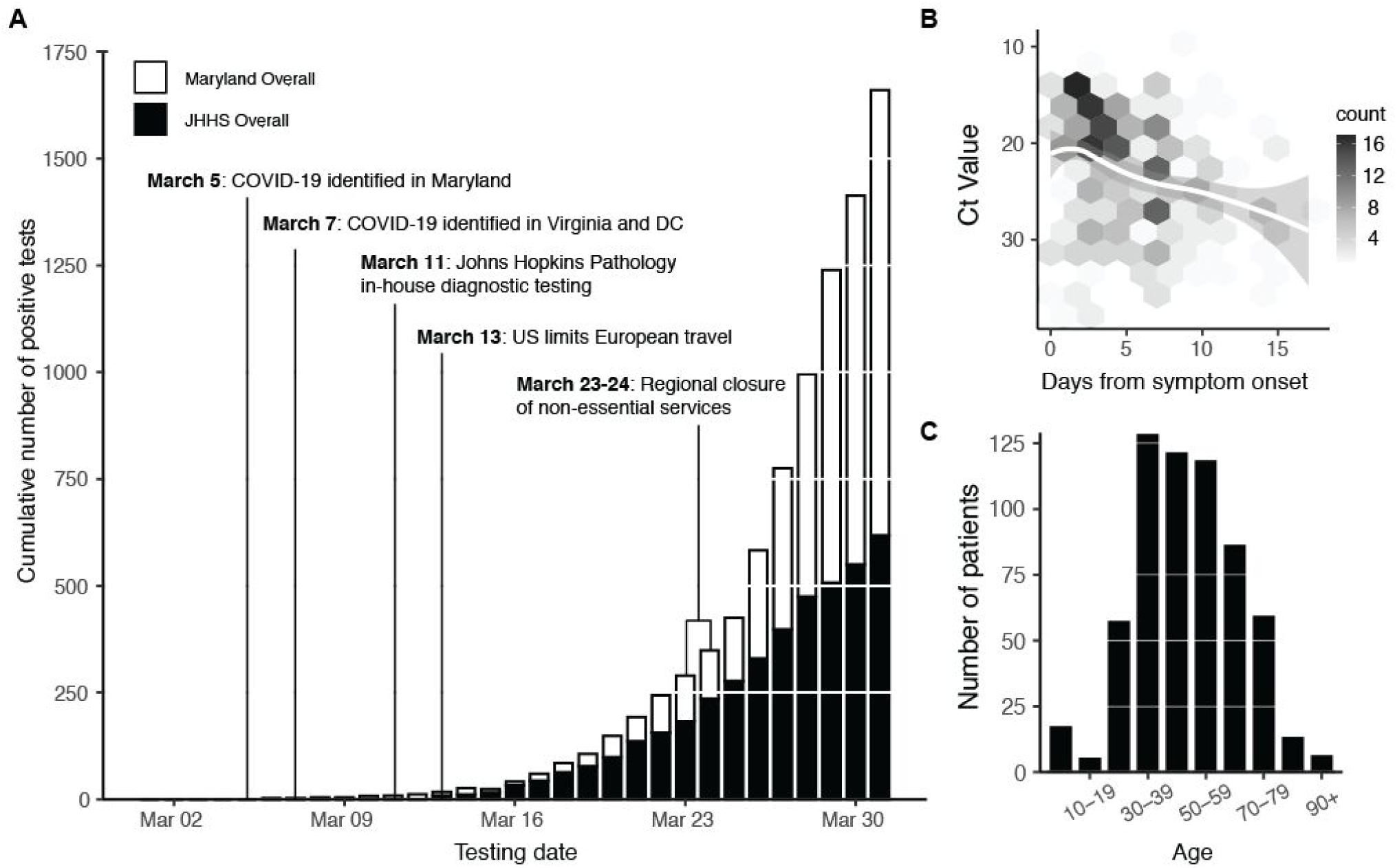
COVID-19 Diagnostic Response During Initial SARS-CoV-2 Surveillance in the Johns Hopkins Health System. **(A)** Cumulative number of positive tests in the state of Maryland (white bars) and within the Johns Hopkins Health System (JHHS; black bars). **(B)** SARS-CoV-2 RT-PCR C_T_ value (S-gene) versus days from patient symptom onset. Data fit with LOESS curve (white regression line). Two outliers (days from onset = 5 weeks, C_T_ value = 30 and days from onset = 28 days, C_T_ value = 31) are not shown. **(C)** Age distribution of SARS-CoV-2 patients within the JHHS.

Molecular diagnosis of SARS-CoV-2 at the Johns Hopkins Hospital Medical Microbiology laboratory began March 11th using the RealStar SARS-CoV-2 RT-PCR kit from Altona Diagnostics, which was granted Food and Drug Administration (FDA) Emergency Use Authorization (EUA) after analytical validation^22^. The Altona test targets both lineage B betacoronavirus E genes and the SARS-CoV-2 S gene. In the last week of March, the laboratory also began the use of the Cepheid Xpert Xpress SARS-CoV-2 assay granted FDA EUA, which targets the E and N2 genes. The laboratory evaluated a total of 5,913 nasopharyngeal swabs and confirmed 620 COVID-19 positive between March 11–March 31, 2020 (10.4% overall positivity rate) (**Table S1**). In total, 603 positive diagnoses were made with the Altona assay in this time period, and 17 positive diagnoses were made with the Cepheid Xpert Xpress SARS-CoV-2 assay. This represented 37.3% of the 1660 confirmed COVID-19 cases in Maryland during this period (**Fig 1A**).

We found that the cycle threshold (C_T_) value of SARS-CoV-2 diagnostic testing was associated with self-reported days from symptom onset (Spearman correlation: ρ=0.35) (**Fig 1B**). The majority of patients diagnosed were older than 30 years (85%; **Fig 1C**). Gender distribution was roughly equal within the patient population (49% female, 51% male; **Table S2**). Patient home residence was captured for 592/620 positive tests, with 82% (n=485) indicating a home address in Maryland, 14% (n=80) in DC, and 1% (n=≤5) in Virginia. The first three digits of patient home zip codes were used to understand the geographic distribution of patients from the National Capital Region (**Fig S1**). The remaining 23 patients (4%) listed primary residences in 11 different states.

### Sequenced Samples and Characteristics of the Virus

We selected 143 samples for whole genome sequencing based on the availability of residual RNA following the diagnostic PCR. Samples were sequenced in two phases, with the first phase enriched for patients admitted to the ICU (55 samples collected March 11-21^st^, containing 14 patients admitted to the ICU), and the second a convenience sample that captured as many samples as possible for sequencing, irrespective of disease severity (88 samples collected March 13-31^st^, containing 10 patients admitted to the ICU).

We performed multiplexed pooled amplicon sequencing as described by the ARTIC network^23^ on Oxford Nanopore instruments (GridION, MinION).We generated 114 complete genomes from the 143 attempted (76%), defined as those that have at least 27,000 (out of 29,903) unambiguous nucleotides (see **Methods**). For validation purposes, a subset of 31 samples were also sequenced on an Illumina MiSeq from the same amplicons (**Table S3**). Sequenced samples ranged in C_T_ from 14 to 38 and samples with C_T_ values less than 30 produced a larger proportion of complete genomes (86%) than samples with higher C_T_ values (39%) (**Fig 2A**). This is consistent with other SARS-CoV-2 sequencing studies using the Oxford Nanopore platform^3,24^. We found no bias in our ability to generate complete genomes across key metadata categories, such as patient age, patient sex, and sample collection date (**Fig 2B-D**).

**Figure 2.**
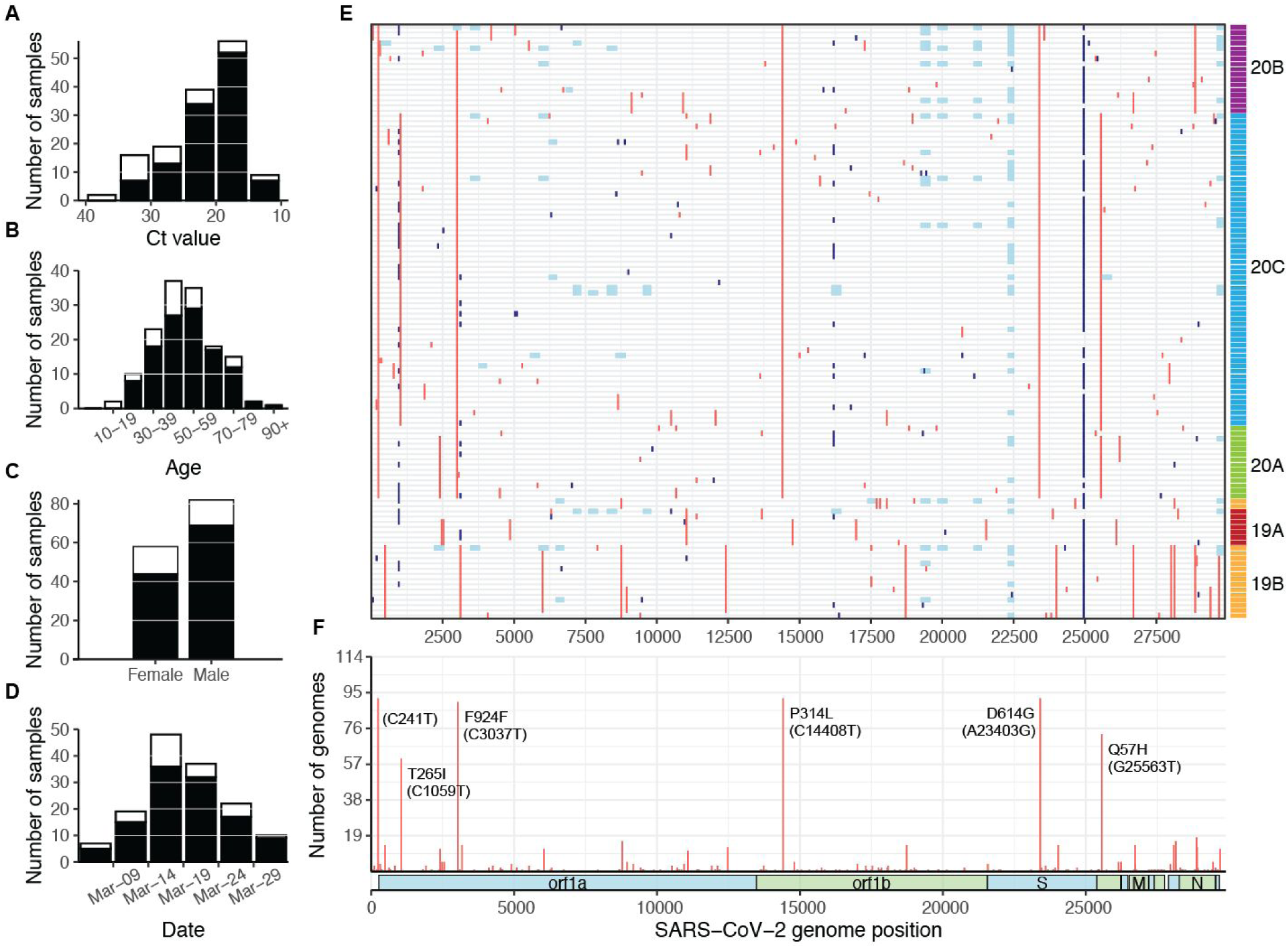
SARS-CoV-2 samples selected for whole genome sequencing. Distribution of (**A**) C_T_ value, (**B**) age, (**C**) sex, and (**D**) collection date for specimens selected for whole genome sequencing (white bars), and specimens that produced complete genomes (black bars). Only specimens with known values are included in each plot. (**E**) Mutations across the SARS-CoV-2 genome in all 114 complete genomes (rows), binned into 60-nucleotide windows. Red = single nucleotide variant, light blue = base masked as N due to amplicon dropout; dark blue = ambiguous base (N) due to variant-calling issues in homopolymer regions. Rows are clustered by Hamming distance between sequences and colored by phylogenetic clades (19A-20C, see **Fig 3**). (**F**) Count of complete genomes (out of 114) with a variant at each site. Key clade-defining mutations are labeled.

The 114 complete genomes correspond to approximately ~18% of JHHS-confirmed cases in March. Incomplete genomes were primarily due to amplicon dropout (26/34 failed samples), leading to stretches of ambiguous basecalls across the genome (**Fig 2E**). Despite these ambiguities, SARS-CoV-2 sequences can be grouped into clusters (or phylogenetic clades) based on a small number of variant sites in high quality regions (**Table S4**).

The 114 sequences were on average 98.6% complete, and we identified a total of 153 unique, unambiguous single nucleotide variants across all sequences (54 synonymous variants, 91 non-synonymous variants, 8 noncoding variants) compared to the Wuhan-Hu-1 SARS-CoV-2 reference genome (accession: MN908947.3) with a range of 2–14 variants per genome. In some samples, we also observed a previously-identified cluster of three nucleotide mutations starting at position 28,881 that always occur together, resulting in two amino acid changes. Within the 114 complete genomes, 20 had one or two mixed sites (25-75% alternate allele frequency), which we replaced with IUPAC ambiguity codes^25^. 12/21 of these non-N ambiguities were at putative C-to-T mutations. We identified 5 clusters of sequences based on their polymorphisms, which correspond to phylogenetic clades (**Fig 2E**). Over half of the sequences (52%) share 5 variants (**Fig 2F**), which corresponds to the phylogenetic clade 20C^15^ (**Table S3**). Within the established clades, we identified 7 small groups of sequences from our National Capital Region dataset (3-14 sequences per group) that share two or more additional single nucleotide variants (**Fig S1, Table S3**).

The amplicon dropout we observed in our incomplete sequences also occurred, to a lesser extent, in our complete sequences — shown as stretches of ambiguous nucleotide calls (**Fig 2E**). In addition to amplicon dropout, evidence of contamination and a high proportion of mixed sites also prevented inclusion of genomes in our final dataset. We observed a small number of sequences (3) that had five or more mixed positions (alternate allele frequency 25-75%, see **Methods**), which required further validation and were not included in the 114 complete genomes used in subsequent analyses. And, despite establishing controlled laboratory workspaces to avoid cross-contamination, we occasionally observed amplification from specific primer sets in negative control samples on a subset of sequencing runs, which caused us to discard the entire sequencing run from analysis. In a small number of cases (5), lack of original specimen availability prevented resequencing and complete genome generation.

To validate our variant calls, we used multiple sequencing approaches, resequencing 31 of the 114 complete genomes on the Illumina platform. We independently called variants from each platform and observed that all Oxford Nanopore variants above 25% allele frequency were also detected using Illumina, and vice versa (**Fig S3A**). We also examined variant sites where resulting in differing consensus nucleotides across the two platforms (due to ambiguity in one consensus and a variant call in the other). Of 280 variant calls made using Oxford Nanopore sequencing, 251 (90%) had the same consensus nucleotide when using Illumina. Of the 29 positions where the consensus calls differed, all but three were due to artificially low Oxford Nanopore allele frequencies at homopolymer sites (with over half occurring at position 3,037) (**Fig S3B, Fig S4A**).

Oxford Nanopore sequencing has known difficulties basecalling in homopolymeric regions^26^, so we were able to correct homopolymer-induced ambiguities in variant sites in our analysis pipeline, thus increasing our confidence in variant calls made on the Oxford Nanopore platform. We also note that the ambiguous basecalls observed in most of our samples at positions 1001, 24981, and 24982 are due to similar issues in homopolymeric regions, but we were only able to systematically correct this issue at variant sites (**Fig 2E**, dark blue). We further validated Oxford Nanopore variants by comparing the results from multiple variant callers. We found Nanopolish to be most reliable for SARS-CoV-2 data, though our pipeline automatically calls variants with multiple callers and reports any discrepancies between them (see **Methods**)^27^.

In addition to basecalling issues in homopolymeric regions, we also observed a strand-specific bias to Oxford Nanopore sequencing at some sites (**Fig S4B**). This bias can occur when the sequence context is more difficult to basecall in one direction, and is corrected in our pipeline by requiring that putative variant alleles are found on reads in both directions.

### Clinical Correlates to Viral Genomics

We performed in-depth chart reviews for all patients with samples selected for sequencing to evaluate potential relationships between the sequence of the virus and disease presentation. These chart reviews captured patient data including comorbidities, symptoms, and disease severity (**Table S2**). We also looked for likely local transmission events, identified by the absence of reported travel in the three weeks prior to diagnosis, as well as likely travel-related importations from locations with known outbreaks in the same time period. In total, 32 (22%) had potential travel exposure from locations with early outbreaks, including England, California, Colorado, New York, and Idaho (Travel history; **Fig 3B**), and 66 (46%) of patients reported having been exposed in a high risk scenario (Known COVID contact; **Fig 3B**). The 111 (78%) individuals that contracted the virus without reported travel history suggest that community transmission was occuring at this early stage of the pandemic.

**Figure 3:**
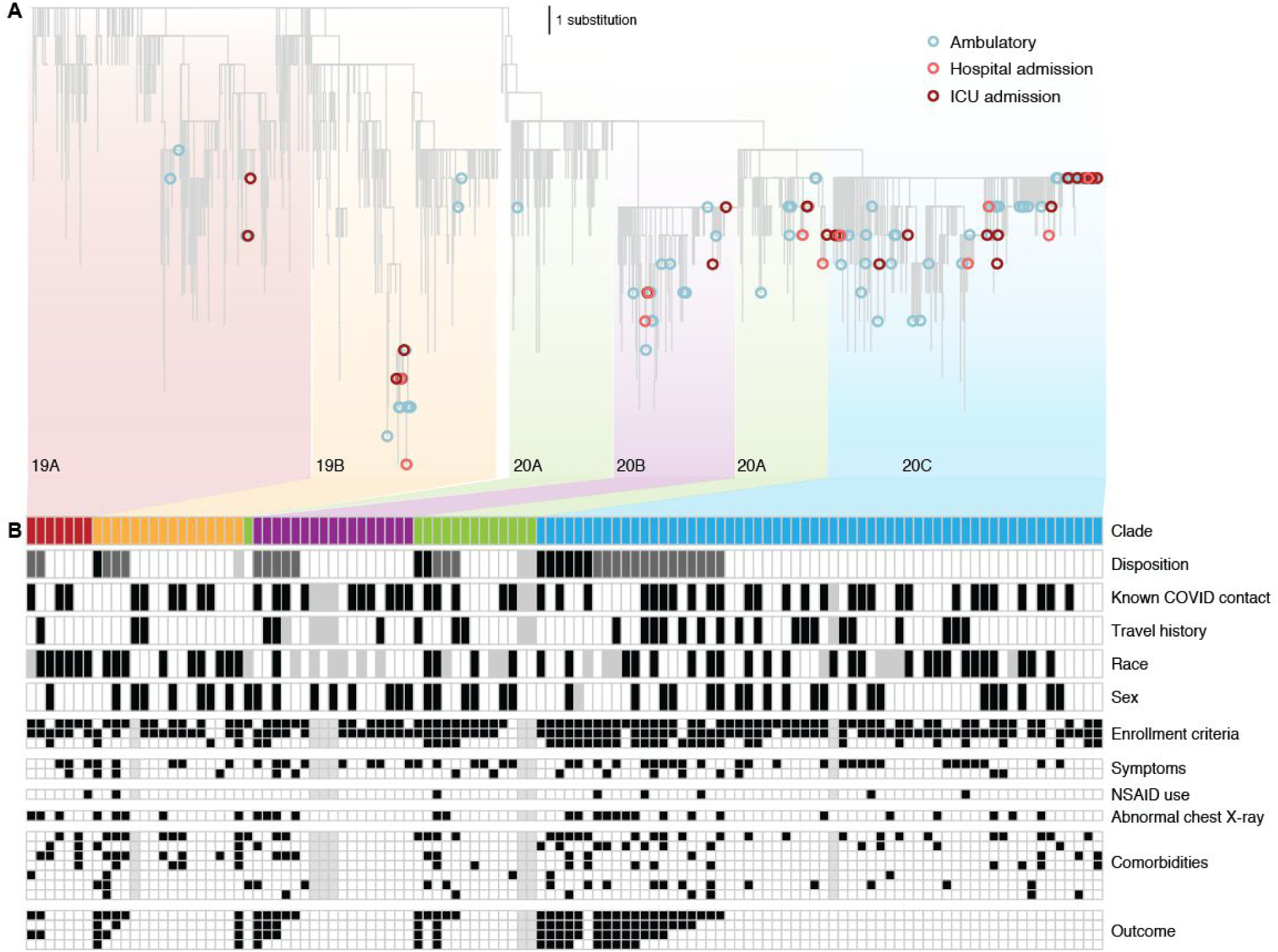
JHHS sequences and patient outcome. (**A**) Maximum likelihood tree of subsampled SARS-CoV-2 global dataset and all 114 sequences generated in this study. Ambulatory (blue) includes all patients with no known admission to the hospital. Hospital admission (light red) includes admitted patients with no known admission to the intensive care unit (ICU), including patients administered oxygen. (**B**) Clinical metadata and virus clade. Each column is one of the 114 patients with virus sequenced in this study, and columns are grouped by disposition within each clade. Unless otherwise specified: black = yes, white = no, gray = unknown. Disposition: black = still in hospital or deceased as of 15-May-2020, dark gray = discharged, white = never admitted. Race: black = Black, white = White, gray = other. Sex: black = female, white = male. Enrollment criteria (top down): Fever, cough, shortness of breath. Symptoms (top down): body ache, GI. Comorbidities (top down): Cardiac disease, Lung disease, diabetes, obese, alcohol, history of smoking (current and former smokers), immunocompromised. Outcome (top down): hospital admission, supplementary oxygen, ICU admission, ventilator administration.

Similar to larger studies^24^, we observed a broad distribution of patient outcomes across the full diversity of SARS-CoV-2 mutations. We observed that severe cases, defined as admission to the ICU (including patients requiring ventilator support), had viral genomes belonging to all five major phylogenetic clades (**Fig 3A**). Similarly, patient phenotypes including sex, race, recent travel, symptoms, and comorbidities were represented across all five major phylogenetic clades, suggesting that susceptbility was independent of clade (**Fig 3B**).

The widely-examined mutation in the viral spike protein (D614G)^28-30^ is one of the key mutations differentiating the 19 and 20 clades. Notably, we see severe cases in both of these clades, though our dataset is underpowered to show significant correlations between viral genome mutations and disease severity. The diversity of virus genetics, clinical symptoms, and patient outcomes suggests that viral mutations are not the main driver of clinical presentation^31,32^.

### Evaluation of Regional and Global SARS-CoV-2 Genetic Diversity

We compared sequences from the National Capital Region to others from around the world to better understand how the virus entered and spread within the region early in the outbreak. We performed phylogenetic analysis using JHHS-generated sequences and a globally-representative reference dataset containing all published sequences collected in Maryland, DC, and Virginia through the end of March 2020 (**Fig 4A**). We see that all major viral clades were represented in the region during this period, suggesting five or more separate introductions, though most sequences belong to clade 20C. Within each clade, we see groups of highly similar or even identical sequences (**Fig 2E**), suggesting that community transmission followed the initial introductions. We also note the presence of a 19B sub-clade containing especially divergent sequences from Maryland and elsewhere. Further studies will be required to understand if the mutation rate is higher in the 19B sub-clade, or if the long branch length to this group is due to biased sampling in global sequence databases.

**Figure 4:**
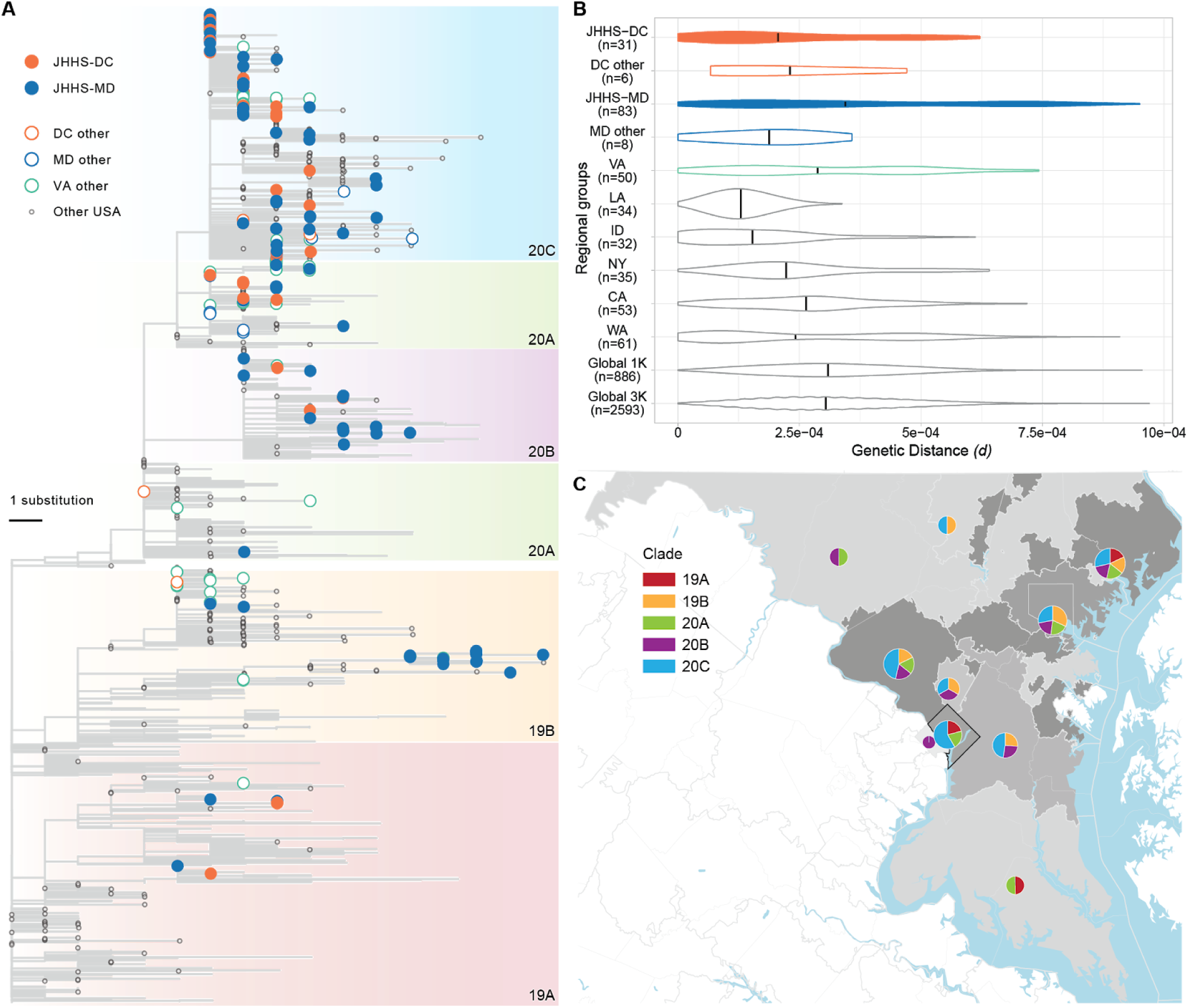
Geographical context of sequences from the National Capital Region. (**A**) Maximum likelihood tree. Filled tips belong to sequences generated in this study. (**B**) Evolutionary divergence in geographic groups. Violin plots represent the distribution of pairwise genetic distances between all sequences for samples collected in each listed geographic group. Abbreviations: Maryland, MD; Virginia, VA; District of Columbia, DC, Washington, WA; California, CA; Idaho, ID; Louisiana, LA; New York, NY. Colors are as in (A), with filled violins containing sequences from this study. Black vertical lines depict the mean pairwise genetic distance between all samples in each regional group. (**C**) Map of the National Capital Region.

We also looked specifically at viral genetic diversity within the National Capital Region, as compared to the total genetic diversity observed in other regions in the US and around the world. We found that the average pairwise viral diversity observed between sequences in the JHHS Maryland dataset is comparable to that of global sequences, concordant with our observation that regional sequences belong to all five major clades (**Fig 4B, Fig S5**). This observation may reflect the national and international connectivity of the entire National Capital Region, as well as the travel patterns of individuals in this region, which includes two major metropolitan areas and three international airports.

We also observed higher mean pairwise genetic distance in the JHHS dataset from Maryland (JHHS-MD) than other data from this state (MD other). Other published sequences from Maryland were submitted by public health laboratories, and lower genetic distances may reflect sequencing of connected clusters of cases during outbreak investigation. The average pairwise genetic distance is lower in other parts of the region (DC and Virginia) than in Maryland, though it is clear that there are sequences from multiple clades present in both locations. It is unclear if the lower observed average distance in both JHHS and other sequences from DC is a reflection of movement patterns of individuals within that nearby area or simply lower sample size (only 32 sequences from DC are in our dataset, compared to 82 total sequences from Maryland). Published sequences collected in Virginia suggest its genetic diversity falls between that of Maryland and DC, but Virginia is a large state, and without more detailed location information for these sequences it is difficult to determine if the sequences truly represent the diversity circulating in the state as a whole (**Fig 4B, Fig S5**).

We also analyzed datasets containing sequences from other US states that experienced outbreaks early in the epidemic. We see that the distribution of viral diversity in the National Capital Region looks very different from diversity in states such as Louisiana and Idaho, which show very low mean genetic diversity during the early epidemic. This could be due to sampling of specific clusters of cases as described above (e.g., cases from the ski resort outbreak in Idaho ^33^), or could occur if early cases in these states were seeded by a single source. This stands in contrast to the outbreaks in Washington and New York, for which sequence data clearly shows multiple introductions ^5^. As expected, the diversity of the viral population (the full distribution of pairwise genetic distances) in New York appears to be similar to that of the National Capital Region, as the outbreaks in both regions were seeded multiple times and contain sequences predominantly belonging to the 2020 (20A, 20B, 20C) clades (**Fig 4A-B, Fig S5**).

Finally, we examined viral genetic diversity within and across the National Capital Region, separating this area into sub-regions defined by the first 3 digits of their ZIP code (ZIP3 area). We found that three ZIP3 locations with the highest number of cases in Maryland and DC each had sequences from 4 or 5 of the major clades, and that all but one ZIP3 location had sequences from more than one clade (**Fig 4C**). We do not observe distinct segregation of viral lineages to particular locations within the region, which highlights both the rapid spread of the virus early in the pandemic and the interconnectedness of this region.

## Discussion

Our genomic dataset from the National Capital Region revealed diversity rivaling that of the worldwide phylogeny, even in an early phase of the epidemic. Sequences from the region belonged to all five major phylogenetic clades, suggesting multiple and diverse introductions from regional or international locations. We also observe minimal diversity within each of these specific clades, suggesting transmission of the virus within local communities after an initial introduction. This pattern of diversity highlights the connectedness of the region to both the national and global epidemic, and the challenges that would likely confront any control strategy predicated on low rates of introduction.

The diversity we observe within the region is also visible on smaller geographic scales, with multiple clades represented within each ZIP3 region in Maryland and in DC. This suggests significant movement of viral lineages within the National Capital Region before regional closures were implemented at the end of March, likely due to local transport and a large number of commuters between Maryland and DC. Further research on more recent COVID-19 cases will be needed to understand how national, state and city-based regulations limiting travel and implementing physical distancing recommendations affected these patterns of spread, as well as the impact of subsequent easing of these restrictions.

The diversity of sequences within this region, combined with detailed clinical metadata obtained through the Johns Hopkins Health System, allowed us to explore the relationship between the SARS-CoV-2 virus and patient presentation and outcome. Specifically, we looked for viral genotypes that demonstrated a connection to disease severity, comorbidities, or patient demographics such as gender and race. We found no clear correlation, but were limited by sample size. It will be important to continue to analyze genomic data alongside clinical metadata as the number of available viral genomes increases to look for potentially subtle associations between the viral genome and patient characteristics^32^.

The analyses described above rely on complete and accurate SARS-CoV-2 sequences. We used a tiled amplicon sequencing approach on the Oxford Nanopore platform to generate sequencing data and found that we were able to achieve complete genomes for a substantial portion of samples attempted. As in previous studies ^24^, we found that samples with higher virus titer (low C_T_ value) more reliably produce complete genomes, and these values can be used to triage samples for sequencing when resources are limited. We also observed some correlation between days from symptom onset and C_T_ value, suggesting that epidemiological surveillance may be most effective if patients are captured early in their course of infection.

We also performed validation on our sequences by using multiple variant callers to detect variants, and by sequencing a subset of samples on the Illumina platform. We found that amplicon-based sequencing with Oxford Nanopore generates correct consensus genome sequences (as compared to Illumina sequences), but with some added ambiguities in specific problematic regions, especially homopolymers. We have developed a pipeline that corrects and flags these issues, and it is our hope that highlighting them in this paper contributes to the overall quality of SARS-CoV-2 sequences generated with this widely-used platform that enables rapid sequencing in a variety of settings.

Moving forward, the pipelines established here will be critical to using genomic surveillance to inform the COVID-19 public health response. When confronting a new disease, the first genomes are the hardest to generate, as they require establishment of laboratory protocols and bioinformatic pipelines that can provide accurate and timely information. Both at Johns Hopkins and elsewhere, this has occurred in record time during the COVID-19 pandemic; and the methods and results presented here will serve as the foundation of continued molecular surveillance of SARS-CoV-2 within the JHHS. This ongoing work should allow us to answer critical questions not only about the evolution of the virus, but on the fundamental mechanisms by which control measures affected its epidemic spread. These efforts complement the information provided by the rapidly growing public databases of SARS-COV-2 sequences by focusing collection of genomic data in settings where we can access extensive current and past clinical data to investigate fundamental questions about this evolving virus’s changing relationship with human health.

## Data Availability

Raw nanopore and Illumina data are deposited at SRA (Bioproject PRJNA629390). Consensus sequences are deposited at GISAID and Genbank (MT509452-MT509493, MT646048-MT646120)

## Acknowledgements

This study was made possible by the tireless efforts of the Johns Hopkins clinical staff and the clinical virology laboratory. We thank Joshua L. Cherry, Kianna Blount, Brian Merritt, Kristina Zudock, Ariel Gershman, Timothy Gilpatrick, and Steven Salzberg for technical support and discussion. We acknowledge the authors and laboratories responsible for generating and submitting sequences into GISAID’s EpiFlu Database. A full table of sequence authors is available in the supplemental information. This work was supported by the Johns Hopkins University President’s Fund Research Response (P.M.T., S.C.R., W.T.), the Johns Hopkins University Applied Physics Laboratory Internal Research and Development (P.M.T.), The Johns Hopkins Hospital (S.W. and J.L.), the Johns Hopkins Department of Pathology, NIH awards U41HG006620 (M.C.S.) and R01AI134384 (M.C.S.), and NSF award DBI-1627442 (M.C.S.). This research makes use of the SciServer science platform (www.sciserver.org). SciServer is a collaborative research environment for large-scale data-driven science. It is being developed at, and administered by, the Institute for Data Intensive Engineering and Science at Johns Hopkins University. SciServer is funded by the National Science Foundation through the Data Infrastructure Building Blocks (DIBBs) program and others, as well as by the Alfred P. Sloan Foundation and the Gordon and Betty Moore Foundation. The opinions expressed in this article are those of the authors and do not reflect the view of the National Institutes of Health, the Department of Health and Human Services, or the United States government.

## Materials and Methods

### Specimens and patient data

Clinical specimens used for genetic characterization were remnant nasopharyngeal swabs available at the completion of standard of care testing at the Johns Hopkins Hospital clinical virology laboratory. In total, 143 samples were selected for analysis based on their distribution throughout March 2020 and representation of the range of disease severity observed during this period. During this period, automated patient metadata extraction was limited to the date a sample was confirmed positive; all other data required patient chart reviews. Samples were sequenced in two phases, with the first phase enriched for patients admitted to the ICU (14/55 samples collected March 11-21^st^), and the second a convenience sample that captured as many samples as possible for sequencing, irrespective of disease severity or ICU admission (10/88 samples collected March 13-31^st^). All research was conducted under Johns Hopkins IRB protocol IRB00221396.

### Clinical data analysis

Data including patient demographics, symptoms, comorbidities, COVID-19 exposure, recent travel history, and results of chest imaging at presentation were abstracted from the electronic medical record (EMR). COVID-19 treatment (medication, supplemental oxygen and invasive mechanical ventilation) and outcomes (home observation without inpatient admission, discharge after admission, ongoing admission, and death) were also abstracted from the EMR. Race as self-reported by the patient and documented in the EMR was collected in pre-specified categories. Patients who reported i) contact with an individual known to be COVID 19 infected or ii) high risk exposure (e.g., healthcare worker) were classified as COVID-19 exposed. Comorbidities were assessed based on diagnoses in the EMR (i.e. Diabetes, Obesity or Alcohol Use Disorder) and further categorized for lung disease (e.g. asthma, COPD), cardiac disease (e.g. valvular heart disease, arrhythmias, hypertension) and immunocompromised (e.g., HIV positive, hematologic malignancy, solid organ transplant).

### Nucleic acid extraction

Automated nucleic acid extraction was performed using either the NucliSENS easyMag or eMAG instruments (bioMérieux) using software version 2.1.0.1. Two mL of easyMag or eMAG lysis buffer was added to 500μL of aliquoted viral transport media in a biosafety cabinet in either BSL-3 facility or BSL-2 using BSL-3 biosafety measures. Specimens were incubated for ten minutes in the lysis buffer prior to automated nucleic acid extraction following the off-board lysis bioMérieux protocol, with an RNA elution volume of 50uL.

### Diagnostic RT-PCR

The Altona Diagnostics RealStar® SARS-CoV-2 RT-PCR Kit 1.0 (Hamburg, Germany) was the primary assay used for molecular diagnosis. A subset of SARS-CoV-2 positives were identified using the Cepheid Xpert Xpress SARS-CoV-2 GeneXpert platform per manufacturer instructions. All samples were processed within 24 hours of collection.

The RealStar® SARS-CoV-2 RT-PCR Kit 1.0 total reaction volume was 30μL (10μL extracted sample and 20μL Master Mix). The kit contains two premade master mixes, A and B, which contain PCR buffer, magnesium salt, primers and probes, reverse transcriptase, and DNA polymerase. The detectors used are Cy5 (SARS-CoV-2; S gene), FAM (B-βCoV; E gene) and JOE (Internal Control). C_T_ values for the S gene target (Cy5) are reported in **Table S2**. Taqman RT- PCR was performed using the Prism 7500 Sequence Detection System (Applied Biosystems, Foster City, CA) at the following cycling conditions: 1 cycle at 55.0°C for 20 minutes, 1 cycle at 95.0 °C for 2 minutes and 45 cycles at 95.0°C for 15 seconds, 55.0°C for 45 seconds then 72.0 °C for 15 seconds. Validation of this assay was performed as described in ^22^.

Testing on the Cepheid Xpert Xpress SARS-CoV-2 GeneXpert platform was performed in accordance with manufacturers instructions ^34^.

### Genome sequencing with ARTIC tiled amplicons

Whole genome amplification of the SARS-CoV-2 genome was performed using the ARTIC network protocol with the V3 primer set ^23^. Briefly, cDNA was generated from previously extracted RNA remaining after the initial diagnostic RT-PCR assay. Unlike the written protocol, no sample dilution was performed to normalize samples by C_T_ value ranges, as this data was often incomplete at the time of sample processing. A two-step reverse transcriptase PCR was performed using random hexamer cDNA synthesis using SuperScript IV (Thermo Fisher, 18091), followed by multiplexed PCR in two non-overlapping pools using Q5 DNA polymerase (New England Biolabs, M0491). For Oxford Nanopore sequencing, amplicon pools were indexed using the Native Barcoding reagent set (Oxford Nanopore, EXP-NBD104). Indexed sample sets of 11 were then pooled, and 20ng of the resulting library was used for sequencing on Oxford Nanopore GridION instruments using R9.4.1 flowcells and high-accuracy basecalling (Guppy v3.5.2).

For Illumina sequencing, the New England Biolabs NEBNext Ultra II DNA Library Prep Kit for Illumina reagent set was used for library generation from the same starting amplicons as used for Oxford Nanopore library preparation. Sequencing adapters were diluted 10 fold to optimize for our input range of 5-100ng of DNA. Adapter-ligated DNA was cleaned up without size selection and underwent 8 cycles of PCR at the amplification step. Samples were sequenced on a MiSeq using a 600bp v3 cartridge.

### Genome assembly and variant validation

Reference-based genome assembly was performed using the ARTIC network bioinformatics pipeline v1.0.0 for COVID-19 (https://artic.network/ncov-2019) with modifications. Briefly, basecalled reads were demultiplexed with Guppy v3.5.2. Reads were mapped to the SARS-CoV-2 reference (GenBank accession MN908947.3) with minimap2 v2.17 ^35^ and coverage was normalized across the genome using a custom normalization pipeline (https://github.com/mkirsche/CoverageNormalization) with coverage_threshold 150 and parameters --even_strand and --qual_sort. Primer binding regions were masked and variant calling was performed with Nanopolish v0.13.2 with a minimum candidate allele frequency of 0.15 ^36^. Consensus genomes were generated with bcftools v1.9 ^37^ by mapping called variants to the reference genome, and all sites with less than 20x coverage were masked as ‘N’.

A custom pipeline was used to validate called variants (https://github.com/timplab/jhu-covid-pipeline). This pipeline made use of the negative control (NTC) on each sequencing run. Amplicon regions with one or more positions with read depth less than two times the 95% quantile of average amplicon depth in the NTC (minimum threshold = 20) were masked as ‘N’, and any variants also present in the NTC were masked unless the coverage at that variant position was more than five times the NTC coverage at that position. Additionally, any sequencing runs with high coverage in the NTC (>50x depth threshold) were ignored and all samples rerun.

Variants in samples or regions without evidence of contamination were validated by two other variant callers: medaka v0.11.5 (implemented by re-running the ARTIC bioinformatics pipeline) and a naïve samtools ^37^ variant caller, both with a minimum candidate allele frequency of 0.15. All positions with variant caller disagreements or high minor allele frequencies (mixed variants; variant allele frequency 25-75%) were manually inspected in Integrated Genome Viewer ^38^. Mixed variants found only on one sequencing strand were ignored (called as reference base), and mixed variants due to deletions in clear homopolymer regions were called as the alternate allele in the consensus genome. For the purposes of creating a consensus genome, candidate variants at <25% allele frequency were called as the reference base, and variants >75% frequency were called as the alternate allele.

When available, Illumina data was used to confirm or invalidate variants with disagreements. The same normalization process was applied to Illumina reads, and variant calling was performed with FreeBayes v0.9.21 ^39^, iVar v1.0 ^40^, and samtools. Mixed variants that could not be confirmed with Illumina data or (in)validated due to strand bias or homopolymer deletions were replaced with ambiguity codes. Final variants were annotated with SnpEff ^41^.

Genomes were considered complete if they had at least 27,000 non-N nucleotide calls (specific IUPAC ambiguity codes such as Y or R contributed to reaching the 27,000 threshold). We also required that the sequence had fewer than 5 mixed variants, as the cause of highly mixed samples (perhaps due to contamination, co-infection, or within-host variation) requires further research.

Genomic analyses were performed on the SciScerver science platform 42. Complete assemblies and all raw sequencing data were submitted to NCBI under bioproject PRJNA650037 (accessions available in **Table S3**).

### Selecting a genomic background dataset

For phylogenetic analyses, full length viral genome sequences with collection dates before April 1, 2020 were downloaded from Genbank ^43^ and GISAID ^18^ on June 3, 2020. Multiple sequence alignment was performed using MAFFT v7.458 ^44^ using parameters --reorder --anysymbol --nomemsave --adjustdirection. Sequences with fewer than 75% unambiguous bases were excluded, as were duplicate sequences defined as having identical nucleotide composition and having been collected on the same date and in the same country. The resulting dataset was trimmed at the 5’ and 3’ ends resulting in a multi-sequence alignment with 29805 nucleotides. This dataset was subjected to multiple iterations of phylogeny reconstruction using IQ-TREE multicore software version v1.6.12 ^45^ with parameters -m GTR+G -nt 50, and exclusion of outlier sequences whose genetic divergence and sampling date were incongruent using TempEst ^46^, resulting in a dataset with 19,565 sequences.

For computational efficiency, we downsampled this dataset homogeneously through time and space, by randomly selecting 7 and 34 sequences per month, to obtain global datasets with 1168 (hereafter referred to as Global 1K) and 3113 (hereafter referred to as Global 3K) sequences from around the world, respectively. We preferentially selected longer sequences with the fewest number of gaps in the 5’ and 3’ ends and those that had complete dates and the fewest number of ambiguous bases. We used the high-performance computational capabilities of the Biowulf Linux cluster at the National Institutes of Health (Bethesda, MD, USA) (http://biowulf.nih.gov) to perform these downsampling analyses. After downsampling, we removed any sequences with fewer than <27,000 unambiguous bases and any remaining sequences deemed GenBank/GISAID duplicates.

In order to study regional epidemics within the United States, sequences from Washington (WA), California (CA), Idaho (ID), Louisiana (LA) and New York (NY) were excluded from the Global 1K and 3K datasets. Data from the National Capital region (Maryland, DC, Virginia) were also removed from these global datasets, resulting in final global datasets of 886 (Global 1K) and 2593 (Global 3K) sequences (**Table S5, Table S6**).

### Comparison of evolutionary divergence

We estimated the evolutionary divergence of several sequence datasets from each of the locations selected for regional analysis. Each regional dataset consisted of sequences removed from the Global 3K data. For Maryland and DC, we supplemented these to include all 114 JHHS sequences. We then subsetted these sequences based on whether they were generated as part of this study (JHHS-MD and JHHS-DC) or from other laboratories (MD other, DC other). For Virginia, we supplemented the removed sequences to include all Virginia sequences in the pre-downsampled global dataset. The final regional datasets were as follows: JHHS-DC (n = 31); DC other (n = 6); DC = JHHS-DC + DC other (n = 37); JHHS-MD (n = 83); MD other (n = 8); MD = JHHS-MD + MD other (n = 91); VA (n = 50); LA (n = 34); ID (n = 32); NY (n = 35); CA (n = 55); WA (n = 53) (**Table S6**).

To estimate evolutionary divergence, we calculated the pairwise divergence (in base substitutions per site) between all pairs of sequences within and between each geographical group. We conducted the analyses through the Molecular Evolutionary Genetics Analysis software version 10 (MEGA X) ^47,48^ and applied the maximum composite likelihood mode ^49^. The rate variation among sites was modeled with a gamma distribution (shape parameter = 4), and the differences in the composition bias among sequences were considered in evolutionary comparisons ^50^. We included 1st+2nd+3rd+non-coding codon positions, and all with less than 50% site coverage, due to alignment gaps, missing data, and ambiguous bases, were eliminated (partial deletion option).

We included the Global 1K and 3K datasets in this analysis to determine the appropriate global reference dataset (out of over 60,000 global SARS-CoV-2 sequences published at the time of analysis) for our phylogenetic analyses. Despite the significant difference in number of sequences, the Global 1K and 3K datasets we tested have comparable mean pairwise genetic distances, indicating that the smaller 1K dataset is representative of global diversity and is an appropriate selection of sequences to use as the background for our phylogenetic inference (**Fig S5**, **Table S5**, **Table S6**).

### Phylogenetic analysis

We used a customized Nextstrain snakemake pipeline ^12^ using augur v9.0.0 on the final dataset, which included the 886 Global 1K dataset plus all removed regional groups (including all 114 JHHS sequences and all published Virginia sequences), resulting in 1279 sequences used in our phylogenetic analysis. We computed the phylogeny with IQ-TREE v1.6.12 ^45,51^ with parameters -me 0.05 -nt 4 -m GTR -n 4. Trees were rooted on the Wuhan-Hu-1 reference genome in FigTree ^52^ and visualized using ggtree ^53^ in R ^54^. Finally, clades were assigned to sequences using Nextstrain ^15^ and Pangolin ^14^, see **Table S3**.

## Supplemental Figures and Tables

**Supplemental Figure 1.**
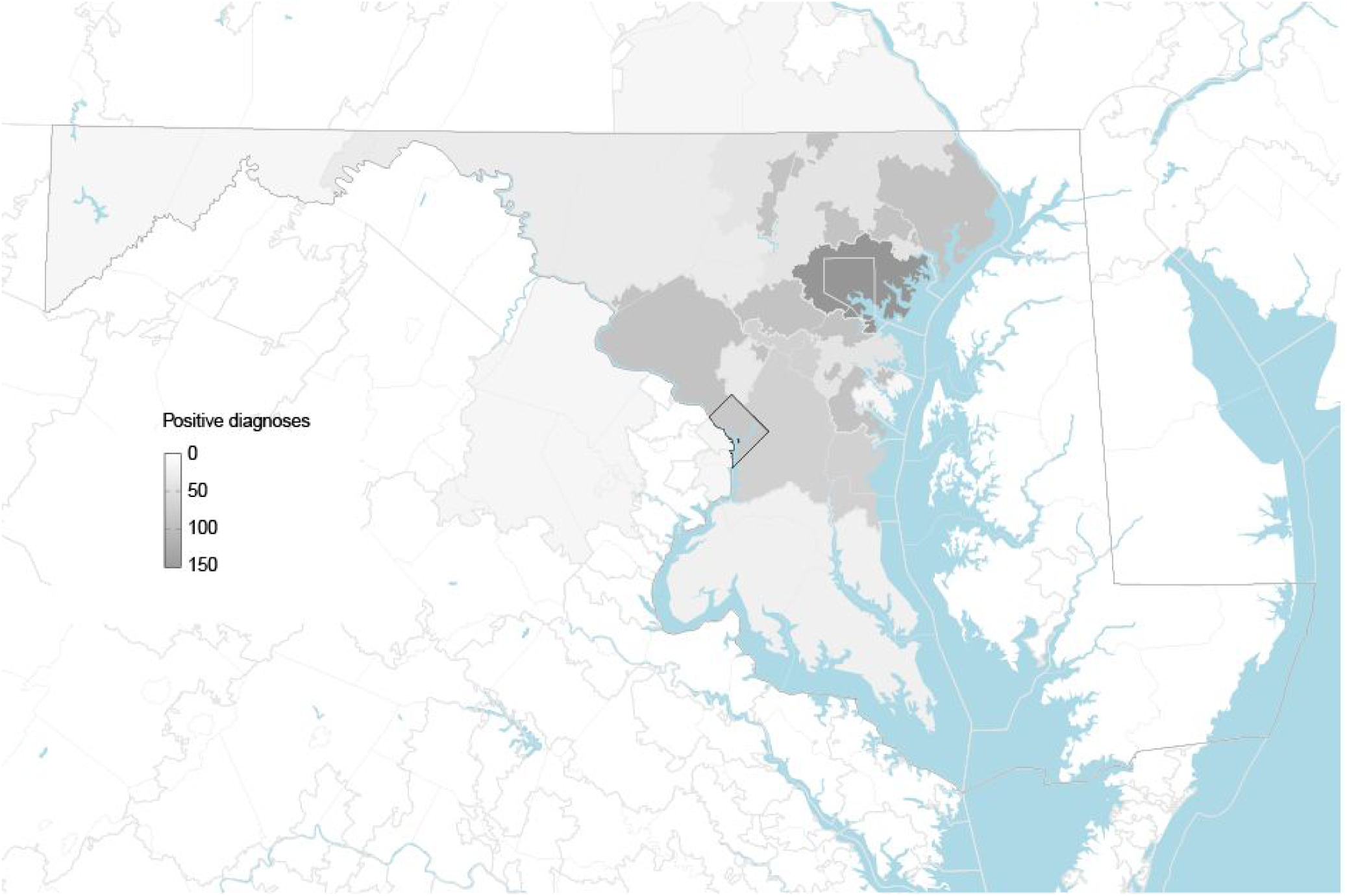
Positive diagnoses by first three digits of zip code. Outlined regions correspond to areas sharing the first three digits of their zip code (Washington DC outlined in black, all others grey). Each region is shaded by the number of patients with positive COVID-19 diagnoses in March 2020 (from the Johns Hopkins Health System) reporting home residence in that region. Regions with 1-5 positive diagnoses are shown as 5.

**Supplemental Figure 2.**
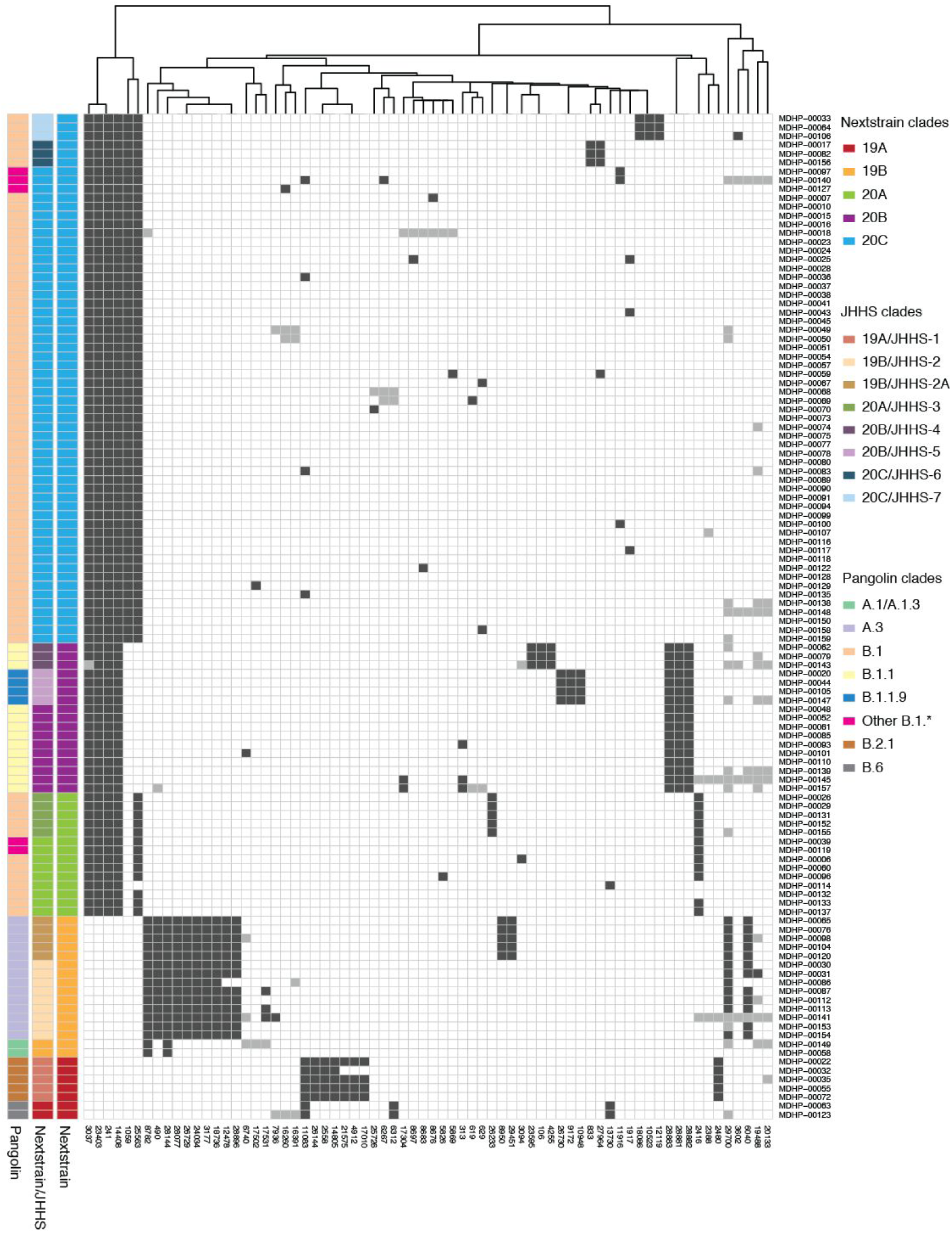
Single nucleotide polymorphisms present in three or more JHHS sequences. Rows = samples sequenced as part of this study (114), sorted by Nextstrain clade. Columns = positions with consensus variants in three or more sequences. Black cells = variant present; grey cells = ambiguity present. Columns are clustered using complete linkage hierarchical clustering. JHHS subclades are additional subclades of Nextstrain clades in which two or more variants are shared between three or more JHHS sequences. Some JHHS-specific clades are captured by Pangolin nomenclature, some are not.

**Supplemental Figure 3.**
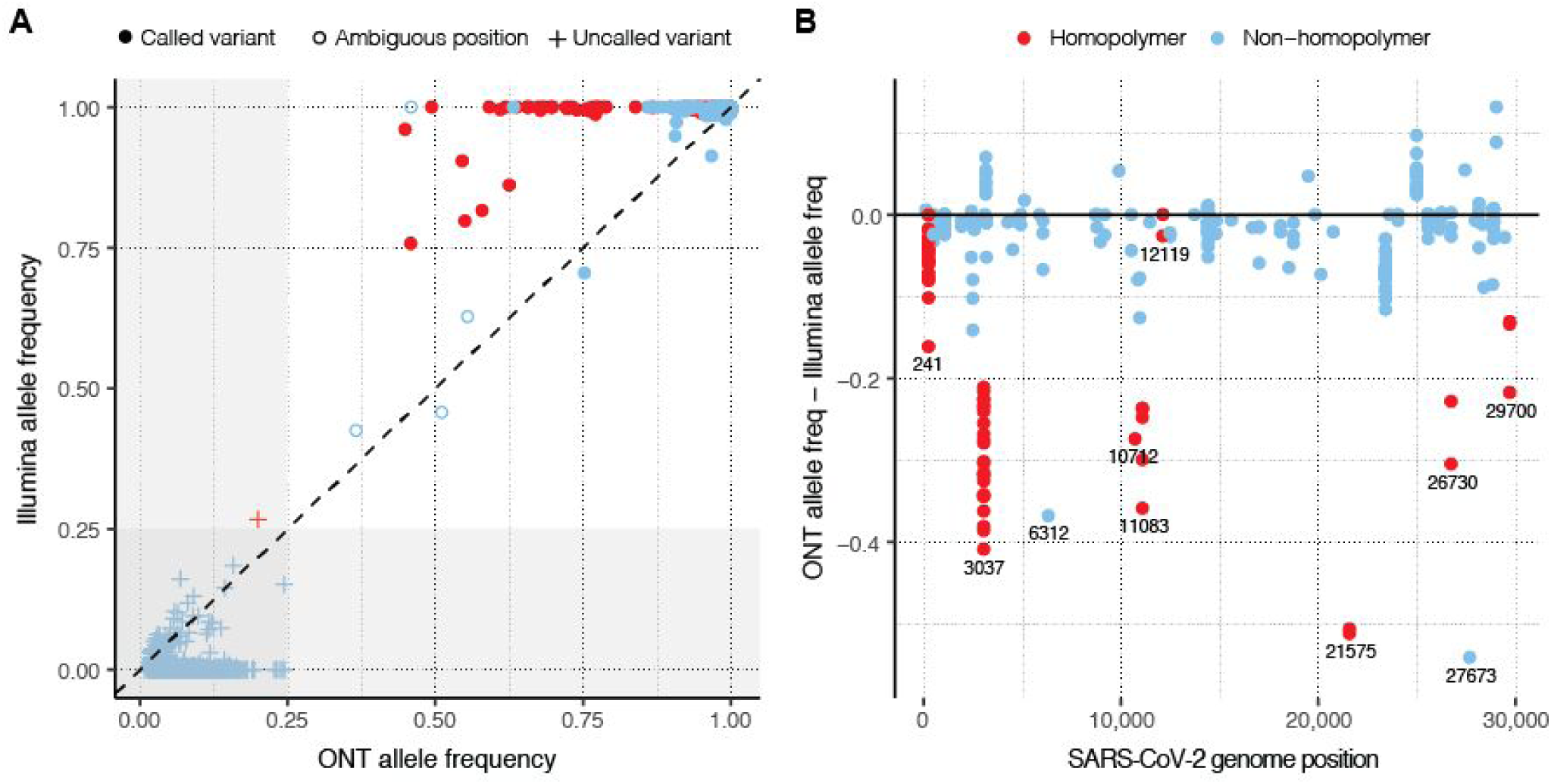
Illumina and Oxford Nanopore allele frequency comparison. (**A**) Variant frequencies from tiled amplicons sequenced on the Oxford Nanopore (ONT) and Illumina platforms. All variants at >0.02 allele frequency are shown. Uncalled variants are those not present in the consensus sequences from these samples. Grey shaded regions represent frequency threshold (25%) used to automatically reject candidate variants. Almost all discrepancies in frequency occur within homopolymer regions (red symbols). (**B**) Difference between allele frequencies across the two sequencing platforms. Position 3,037 (within a T-homopolymer) accounts for most large discrepancies in frequency.

**Supplemental Figure 4.**
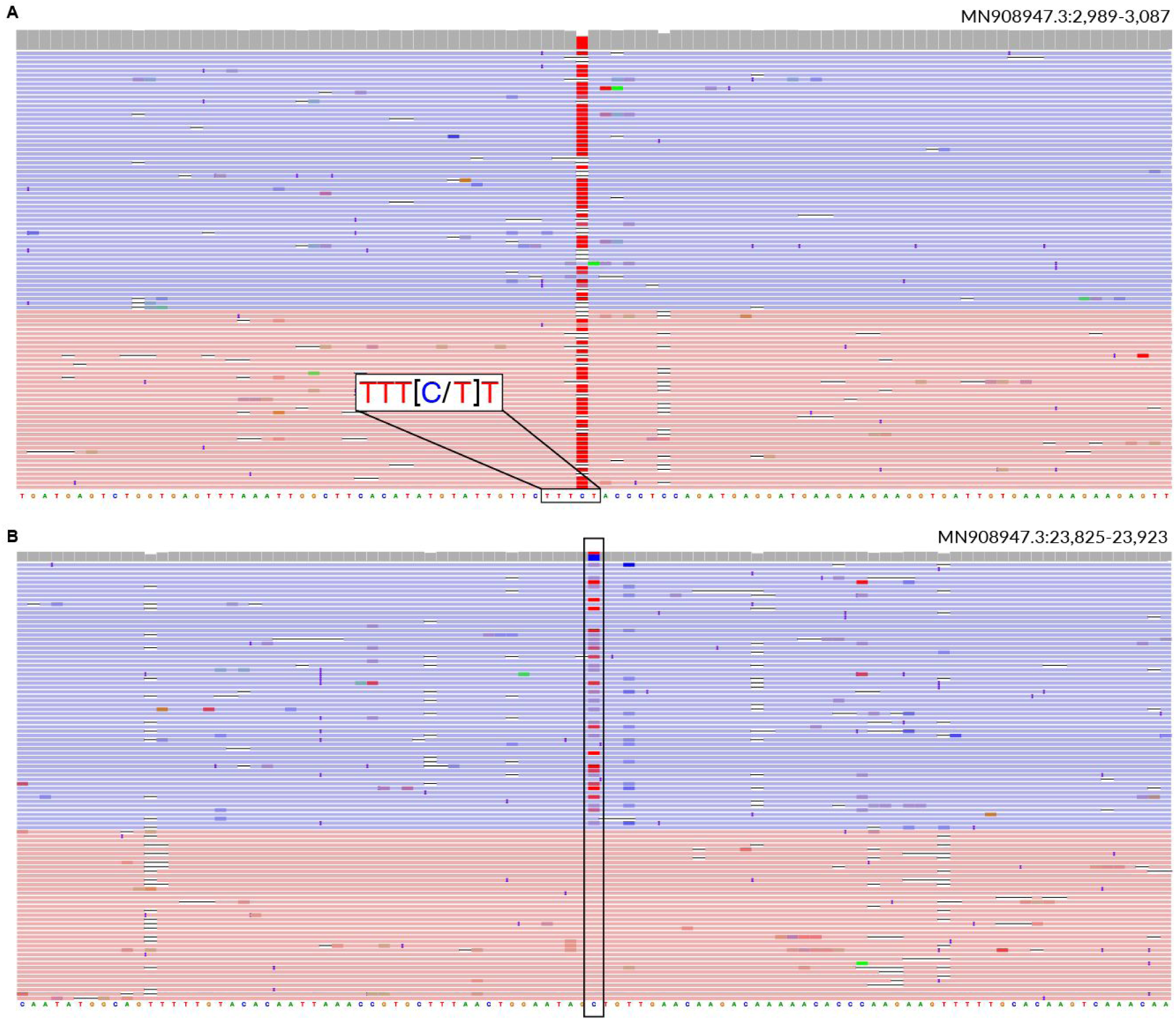
Common Oxford Nanopore sequencing issues. (**A**) Example of a T-homopolymer region in the SARS-CoV-2 genome (sample MDHP-00028). A large number of reads have deletions, resulting in a mixed alternate allele frequency. (**B**) Example of strand bias in Oxford Nanopore sequencing reads (sample MDHP-00028). A mutation only occurs in minus strand reads.

**Supplemental Figure 5.**
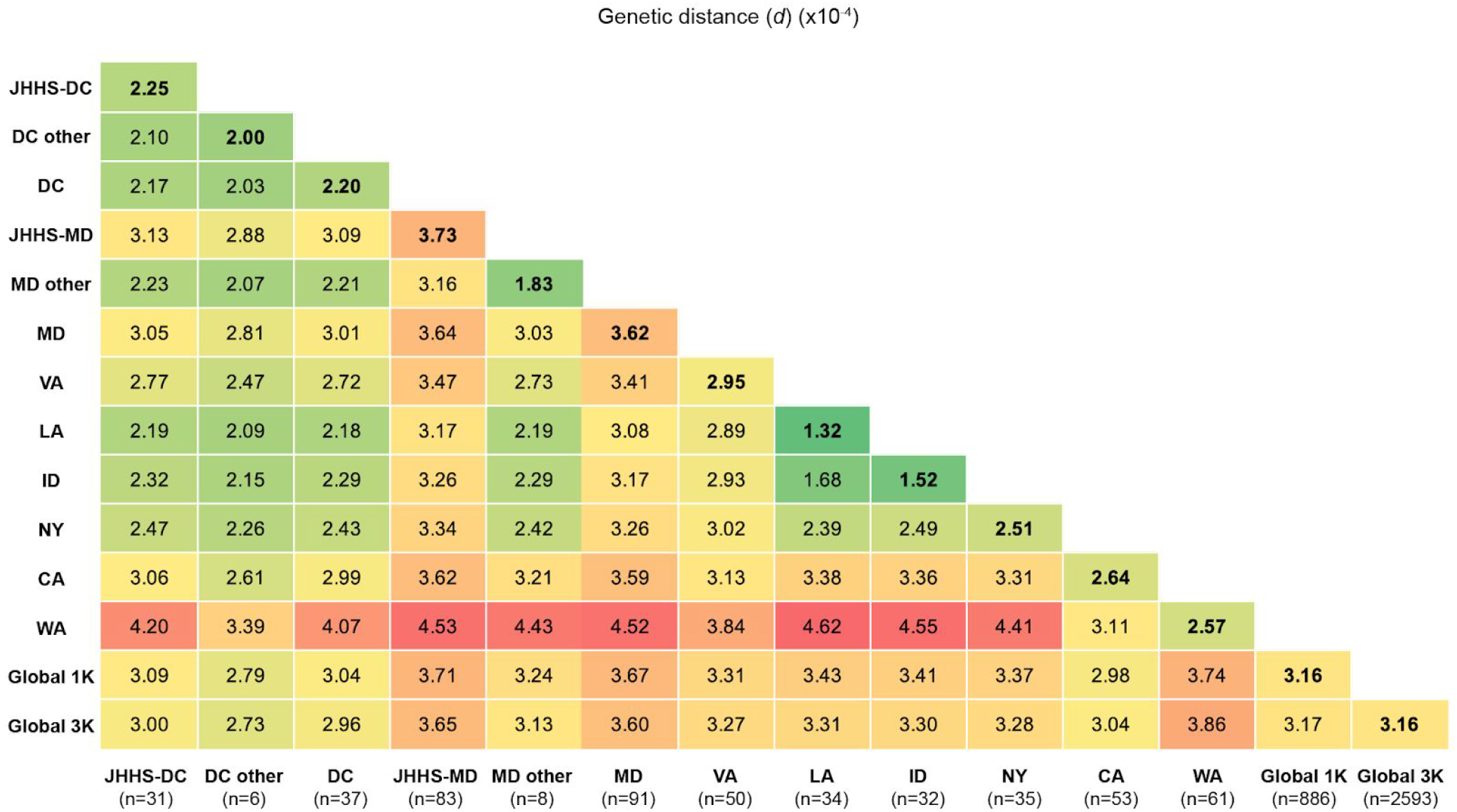
Evolutionary divergence in geographic regions. Pairwise matrix containing average pairwise genetic distances within (bolded in the diagonal) and between sequences from various geographic regions including: the District of Columbia (JHHS-DC + DC other), Maryland (JHHS-MD + MD other), Virginia (VA), Louisiana (LA), Idaho (ID), New York (NY), California (CA), Washington (WA), and two representative global subsamples (Global 1K and 3K). Green-to-red color scale highlights the most (red) and least (green) divergent within or between average distances.

**Supplemental Table 1. Aggregate JHHS diagnostic tests by date**.

**Supplemental Table 2. Aggregate sample metadata**.

**Supplemental Table 3. Sequencing metrics**.

**Supplemental Table 4. Single nucleotide polymorphisms present in JHHS sequences**.

**Supplemental Table 5. Accession numbers of samples used in phylogenetic analysis**.

**Supplemental Table 6. Accession numbers of samples used in genetic distance analysis**.

## References

1 World Health Organization. Coronavirus disease 2019 (COVID-19) Situation Report – 51. 2020; published online March 11. https://www.who.int/docs/default-source/coronaviruse/situation-reports/20200311-sitrep-51-covid-19.pdf?sfvrsn=1ba62e57_4.

2 Dong E, Du H, Gardner L. An interactive web-based dashboard to track COVID-19 in real time. Lancet Infect Dis 2020; 20: 533–4.

3 Lu J, du Plessis L, Liu Z, et al. Genomic Epidemiology of SARS-CoV-2 in Guangdong Province, China. Cell 2020; 181: 997–1003.e9.

4 Bedford T, Greninger AL, Roychoudhury P, et al. Cryptic transmission of SARS-CoV-2 in Washington State. *medRxiv* 2020; published online April 6. DOI:10.1101/2020.04.02.20051417.

5 Gonzalez-Reiche AS, Hernandez MM, Sullivan MJ, et al. Introductions and early spread of SARS-CoV-2 in the New York City area. Science 2020; 369: 297–301.

6 Fauver JR, Petrone ME, Hodcroft EB, et al. Coast-to-Coast Spread of SARS-CoV-2 during the Early Epidemic in the United States. Cell 2020; 181: 990–6.e5.

7 Seemann T, Lane C, Sherry N, et al. Tracking the COVID-19 pandemic in Australia using genomics. Infectious Diseases (except HIV/AIDS). 2020; published online May 16. DOI:10.1101/2020.05.12.20099929.

8 Deng X, Gu W, Federman S, et al. Genomic surveillance reveals multiple introductions of SARS-CoV-2 into Northern California. Science 2020; 369: 582–7.

9 CDC COVID-19 Response Team, Jorden MA, Rudman SL, et al. Evidence for Limited Early Spread of COVID-19 Within the United States, January-February 2020. MMWR Morb Mortal Wkly Rep 2020; 69: 680–4.

10 Stadlbauer D, Tan J, Jiang K, et al. Seroconversion of a city: Longitudinal monitoring of SARS-CoV-2 seroprevalence in New York City. Epidemiology. 2020; published online June 29. DOI: 10.1101/2020.06.28.20142190.

11 Chu HY, Englund JA, Starita LM, et al. Early Detection of Covid-19 through a Citywide Pandemic Surveillance Platform. N Engl J Med 2020; 383: 185–7.

12 Hadfield J, Megill C, Bell SM, et al. Nextstrain: real-time tracking of pathogen evolution. Bioinformatics 2018; 34: 4121–3.

13 Andersen KG, Rambaut A, Lipkin WI, Holmes EC, Garry RF. The proximal origin of SARS-CoV-2. Nat Med 2020; 26: 450–2.

14 Rambaut A, Holmes EC, Hill V, et al. A dynamic nomenclature proposal for SARS-CoV-2 to assist genomic epidemiology. bioRxiv. 2020;: 2020.04.17.046086.

15 Emma B Hodcroft, James Hadfield, Richard A Neher, Trevor Bedford. Year-letter Genetic Clade Naming for SARS-CoV-2 on Nextstain.org. 2020; published online June 6. https://nextstrain.org/blog/2020-06-02-SARSCoV2-clade-naming (accessed Aug 2, 2020).

16 Alm E, Broberg EK, Connor T, et al. Geographical and temporal distribution of SARS-CoV-2 clades in the WHO European Region, January to June 2020. Eurosurveillance 2020; 25: 2001410.

17 Beach MC, Lederman HM, Singleton M, et al. Desperate Times: Protecting the Public From Research Without Consent or Oversight During Public Health Emergencies. Ann Intern Med 2020; published online July 27. DOI:10.7326/M20-4631.

18 Elbe S, Buckland-Merrett G. Data, disease and diplomacy: GISAID’s innovative contribution to global health. Glob Chall 2017; 1: 33–46.

19 Quick J, Loman NJ, Duraffour S, et al. Real-time, portable genome sequencing for Ebola surveillance. Nature 2016; 530: 228–32.

20 Faria NR, Sabino EC, Nunes MRT, Alcantara LCJ, Loman NJ, Pybus OG. Mobile real-time surveillance of Zika virus in Brazil. Genome Med 2016; 8: 97.

21 Governor Larry Hogan – Official Website for the Governor of Maryland. MD State of Emergency Declaration. https://governor.maryland.gov/2020/03/05/governor-larry-hogan-declares-state-of-emergency-expands-statewide-response-to-novel-coronavirus/ (accessed Aug 11, 2020).

22 Uhteg K, Jarrett J, Richards M, et al. Comparing the analytical performance of three SARS-CoV-2 molecular diagnostic assays. J Clin Virol 2020; 127: 104384.

23 Quick J. nCoV-2019 sequencing protocol v1 (protocols.io.bbmuik6w). DOI:10.17504/protocols.io.bbmuik6w.

24 Meredith LW, Hamilton WL, Warne B, et al. Rapid implementation of real-time SARS-CoV-2 sequencing to investigate healthcare-associated COVID-19 infections. Infectious Diseases (except HIV/AIDS). 2020; published online July 14. DOI:10.1016/S1473-3099(20)30562-4.

25 Cornish-Bowden A. Nomenclature for incompletely specified bases in nucleic acid sequences: recommendations 1984. Nucleic Acids Res 1985; 13: 3021–30.

26 Wick RR, Judd LM, Holt KE. Performance of neural network basecalling tools for Oxford Nanopore sequencing. Genome Biol 2019; 20: 129.

27 Simpson J. nanopolish. Github https://github.com/jts/nanopolish (accessed May 21, 2020).

28 Korber B, Fischer WM, Gnanakaran S, et al. Tracking Changes in SARS-CoV-2 Spike: Evidence that D614G Increases Infectivity of the COVID-19 Virus. Cell 2020; published online July 3. DOI:10.1016/j.cell.2020.06.043.

29 Yurkovetskiy L, Wang X, Pascal KE, et al. Structural and Functional Analysis of the D614G SARS-CoV-2 Spike Protein Variant. DOI:10.1101/2020.07.04.187757.

30 Zhang L, Jackson CB, Mou H, et al. The D614G mutation in the SARS-CoV-2 spike protein reduces S1 shedding and increases infectivity. *bioRxiv* 2020; published online June 12. DOI:10.1101/2020.06.12.148726.

31 de Souza WM, Buss LF, Candido D da S, et al. Epidemiological and clinical characteristics of the COVID-19 epidemic in Brazil. Nat Hum Behav 2020; published online July 31. DOI:10.1038/s41562-020-0928-4.

32 Volz EM, Hill V, McCrone JT, et al. Evaluating the effects of SARS-CoV-2 Spike mutation D614G on transmissibility and pathogenicity. Epidemiology. 2020; published online Aug 4. DOI: 10.1101/2020.07.31.20166082.

33 McLaughlin CC, Doll MK, Morrison KT, et al. High Community SARS-CoV-2 Antibody Seroprevalence in a Ski Resort Community, Blaine County, Idaho, US. Preliminary Results. medRxiv 2020; published online July 21. DOI:10.1101/2020.07.19.20157198.

34 Cepheid Xpert Xpress SARS-CoV-2 Protocol. 2020 https://www.fda.gov/media/136314/download.

35 Li H. Minimap2: pairwise alignment for nucleotide sequences. Bioinformatics 2018; 34: 3094–100.

36 Loman NJ, Quick J, Simpson JT. A complete bacterial genome assembled de novo using only nanopore sequencing data. Nat Methods 2015; 12: 733–5.

37 Li H. A statistical framework for SNP calling, mutation discovery, association mapping and population genetical parameter estimation from sequencing data. Bioinformatics 2011; 27: 2987–93.

38 Robinson JT, Thorvaldsdottir H, Winckler W, et al. Integrative genomics viewer. Nat Biotechnol 2011; 29: 24–6.

39 Garrison E, Marth G. Haplotype-based variant detection from short-read sequencing. arXiv [q-bio.GN]. 2012; published online July 17. http://arxiv.org/abs/1207.3907.

40 Grubaugh ND, Gangavarapu K, Quick J, et al. An amplicon-based sequencing framework for accurately measuring intrahost virus diversity using PrimalSeq and iVar. Genome Biol 2019; 20: 8.

41 Cingolani P, Platts A, Wang LL, et al. A program for annotating and predicting the effects of single nucleotide polymorphisms, SnpEff: SNPs in the genome of Drosophila melanogaster strain w1118; iso-2; iso-3. Fly 2012; 6: 80–92.

42 Taghizadeh-Popp M, Kim JW, Lemson G, et al. SciServer: a Science Platform for Astronomy and Beyond. arXiv [astro-ph.IM]. 2020; published online Jan 23. http://arxiv.org/abs/2001.08619.

43 Clark K, Karsch-Mizrachi I, Lipman DJ, Ostell J, Sayers EW. GenBank. Nucleic Acids Res 2016; 44: D67-72.

44 Katoh K, Asimenos G, Toh H. Multiple alignment of DNA sequences with MAFFT. Methods Mol Biol 2009; 537: 39–64.

45 Nguyen L-T, Schmidt HA, von Haeseler A, Minh BQ. IQ-TREE: a fast and effective stochastic algorithm for estimating maximum-likelihood phylogenies. Mol Biol Evol 2015; 32: 268–74.

46 Rambaut A, Lam TT, Max Carvalho L, Pybus OG. Exploring the temporal structure of heterochronous sequences using TempEst (formerly Path-O-Gen). Virus Evol 2016; 2: vew007.

47 Kumar S, Stecher G, Li M, Knyaz C, Tamura K. MEGA X: Molecular Evolutionary Genetics Analysis across Computing Platforms. Mol Biol Evol 2018; 35: 1547–9.

48 Stecher G, Tamura K, Kumar S. Molecular Evolutionary Genetics Analysis (MEGA) for macOS. Mol Biol Evol 2020; 37: 1237–9.

49 Tamura K, Nei M, Kumar S. Prospects for inferring very large phylogenies by using the neighbor-joining method. Proc Natl Acad Sci U S A 2004; 101: 11030–5.

50 Tamura K, Kumar S. Evolutionary distance estimation under heterogeneous substitution pattern among lineages. Mol Biol Evol 2002; 19: 1727–36.

51 Hoang DT, Chernomor O, von Haeseler A, Minh BQ, Vinh LS. UFBoot2: Improving the Ultrafast Bootstrap Approximation. Mol Biol Evol 2018; 35: 518–22.

52 Rambaut A. FigTree. http://tree.bio.ed.ac.uk/software/figtree/ (accessed Aug 2, 2020).

53 Yu G, Smith DK, Zhu H, Guan Y, Lam TT. ggtree: an r package for visualization and annotation of phylogenetic trees with their covariates and other associated data. Methods Ecol Evol 2017; 8: 28–36.

54 R Core Team. R: A Language and Environment for Statistical Computing. 2017. https://www.R-project.org/.

